# Adolescent Dating and HIV Perceptions: A Phenomenological Study in N’Djamena, Chad

**DOI:** 10.1101/2024.03.28.24304942

**Authors:** Esias Bedingar, Ngarossorang Bedingar, Djimet Seli, Christopher Sudfeld

**Author notes:** Corresponding author: Esias Bedingar, 655 Huntington Avenue, Boston, MA 02215, USA. E-mail addresses of authors: NB, DS, CS.

## Abstract

**Introduction:** The study focuses on understanding the complex interplay between dating behaviors and HIV perceptions among adolescents and young people (ages 15-24 years) in Chad. As adolescents and young people are disproportionately affected by the HIV epidemic, this research aims to uncover how cultural and social influences shape young people’s attitudes towards dating and HIV prevention. The significance of this study lies in its potential to inform targeted, culturally sensitive HIV prevention strategies for the youth in Chad.

**Methods:** A qualitative phenomenological approach was adopted, using 4 focus group discussions (n=12 each) with high school students in N’Djamena, Chad. Participants were divided into beneficiary and non-beneficiary groups based on their exposure to a peer education program about HIV. Data analysis was conducted using ATLAS.ti software and the descriptive Colaizzi method, ensuring a comprehensive understanding of participants’ perspectives.

**Results:** Findings revealed significant gender differences in dating motivations and partner selection criteria. While males primarily viewed dating as a means to fulfill sexual desires, females often sought emotional connection and companionship. There was a general lack of awareness about PrEP, with more emphasis on condom use for pregnancy prevention rather than HIV or STI protection. Gender roles played a substantial part in decisions regarding safe sex practices, with a notable discrepancy in shared versus individual responsibility. Additionally, risk behaviors like alcohol use and multiple partnerships were identified as prevalent among youth.

**Conclusions:** The study highlights the need for gender-sensitive educational interventions focusing on the realities of adolescent sexual behavior in Chad. It underscores the disparity in HIV knowledge and prevention awareness among adolescents, pointing out the absence of awareness about PrEP. Future research should emphasize developing HIV prevention strategies that resonate with the unique cultural and social dynamics of Chadian youth, considering their specific dating behaviors and perceptions towards HIV.

## 1. Introduction

Young people ages 15-24 years contribute to almost half of all new HIV infections globally [1]. Approximately, 80% of HIV-positive youth reside in sub-Saharan Africa (SSA) [2]. Chad is a country located in Central Africa with an overall prevalence of 1.0% in 2022 [3]. It is estimated that the HIV prevalence among youth ages 15-24 years is 1.3% for females and 0.8% for males [4]. Notably, within the female youth age group, the prevalence increases with age with a prevalence of 1.2% for age 15-19 years, 1.8% for the 18-19 years, and 2.4% for 23-24 years [4]. Interventions to reduce incidence among adolescents are not a focus in Chad while intervention strategies emphasize preventing mother-to-child transmission [4]. The focus on PMTCT may be due to Chad’s cultural norms, where discussing sexuality remains a sensitive topic, which limits open conversations about reproductive health [5].

In Chad, understanding the dynamics of dating culture is crucial for comprehending the risks and behaviors associated with HIV among youth. The dating scene among Chadian youth, characterized by traditional customs intertwined with modern influences, plays a pivotal role in shaping their perceptions and actions related to HIV risk. However, adolescents face substantial challenges in accessing reliable information and resources to HIV [5]. In this context, peer education in schools has emerged as a potential strategy for behavior change [6]. Peer-led HIV education programs in schools aim to bridge this gap by providing accurate information and fostering supportive environments and these programs can leverage the unique position of peers who can communicate complex health messages in a relatable and understandable manner [7]. In 2012, the “Life Skills and Peer Education” initiative, also known as the BCC Lifeskills Project (2012-2016), was initiated by Blue Cross Chad for at-risk students aged 14-18 in schools [8, 9]. This project’s goal was to enhance understanding and skills for making educated choices regarding substance abuse, alcohol, and the risks linked with HIV. The program included peer educators trained in life skills who covered various subjects, from HIV prevention to transmission, using interactive teaching methods like group activities and role-playing.

From 2012 to 2016, the BCC LBSE was conducted in 15 high schools in N’Djamena and later expanded to 20 schools by 2021. With the desire to inform programmatic strategies for scale-up, we conducted a qualitative study to explore the experiences of the youth in reproductive health and HIV and AIDS. The study also aimed to understand how the dating culture and practices influence adolescents’ knowledge, misconceptions, and prevention strategies regarding HIV. Insights from this study are intended to inform the development of effective HIV prevention and education programs tailored to the unique context of Chadian adolescents.

## 2. Methods

### 2.1. Study setting, design, and participants

This qualitative research study was carried out in N’Djamena, which was purposefully chosen because Blue Cross Chad (BCC) had introduced its program in 20 local high schools. N’Djamena is Chad’s largest and capital city. With a population of 1 538 387 people, N’Djamena is composed of 10 districts [10]. As previously mentioned, the HIV prevalence stood at 1.0% in 2022 [3]. Although there is a 48% decrease in new HIV infections since 2010, adolescents are disproportionately affected by the epidemic [11]. According to UNAIDS, 26.3% of new HIV infections were in adolescents and young people (ages 15-24 years) in 2022 [11]. Prior to this, they represented 28.4% and 30.8% of new HIV infections in 2010 and 2015, respectively [11]. Despite all the progress made, young women continue to suffer disproportionately from the HIV epidemic, more specifically due to the HIV triple threat, which is known as the combination of new HIV infections, sexual and gender-based violence, and adolescent pregnancies [12]. Young women have HIV infection rates four times as high as young men, accounting for 19.5% of all new HIV infections [11].

The research design was based on descriptive phenomenology, adhering to the philosophical principles established by Edmund Husserl [13]. As we were interested in the youth’s experience of HIV supportive norms, the methodology was particularly effective in capturing and understanding the unique perspectives and experiences of the participants [14, 15]. Husserl’s four-step process of descriptive phenomenology – bracketing, intuiting, analyzing, and interpreting – guided the research [16]. The research took careful measures to maintain objectivity, especially in focus group discussions (FGDs), by regularly discussing and setting aside any preconceived notions about youth behavior. This was crucial since the interviewers were part of the same organization that aimed to increase HIV prevention awareness, necessitating a methodological approach that prioritized describing experiences over-interpreting them.

In selecting participants, a purposeful criterion was used, targeting students from both schools that participated in the BCC LBSE program and those that did not, to effectively evaluate the program’s impact. FGDs were further stratified by gender to create a comfortable environment for participants to openly share their views. The criteria for selection included students currently enrolled in grades 9 to 12. For students in participating schools, a minimum of six months’ participation in the program was required to ensure adequate exposure to the intervention. The selection process involved using student lists from school registrars, excluding those with incomplete information. The study formed four groups of 12 participants each. Despite the typical FGD size being 6-8 participants, a larger group size was chosen to enhance the diversity of perspectives and group dynamics. However, due to the exploratory nature of the study, saturation was not reached. To recruit participants, especially those under 18, school officials contacted parents to inform them about the research and its objectives. Parental consent was then obtained either in writing, during a visit to the school for those who agreed over the phone, or through verbal consent if an in-person visit was not feasible. Additionally, the COREQ checklist was employed to ensure thorough and transparent reporting of the qualitative research methods and findings [17], as detailed in the supporting information (S2 Appendix).

### 2.2. Data collection and analysis

The data was collected between October 4^th^ and 6^th^, 2022. In the qualitative component of the study, six staff members from the BCC program were selected as interviewers. These individuals were organized into two groups, each comprising three researchers, with at least one female interviewer in each team. Before commencing the study, they received training in qualitative research methodologies, including interview techniques. The decision to use interviewers who were locally trained and familiar with the community was advantageous for gaining access and acceptance within the community. Although we used interviewers who were part of the same organization implementing BCC LBSE, the interviewers selected were not directly involved in the execution of the program, which ensured an unbiased approach.

FGDs were carried out in French and Arabic using a semi-structured guideline (S1 Appendix). The interviews were conducted in a private and secure location at the BCC headquarters. Each FGD lasted between 90-120 minutes, with the transcription process taking about 3-4 hours per session. The team collectively reviewed each transcript to accurately capture cultural nuances. During the FDGs, participants were encouraged to express their views, perceptions, experiences, and beliefs about sexuality, particularly in the context of HIV transmission and prevention. The sessions began with an overview of the interview’s purpose, assurances of confidentiality, and the collection of either written or oral informed consent from the respondents. All discussions were meticulously recorded, transcribed verbatim, and then translated by the entire team, ensuring that the translations retained the cultural context and nuances.

For data analysis, the team utilized ATLAS.ti version 22, a qualitative analysis software [18]. The descriptive Colaizzi method was used to analyze the data, which emphasized the importance of participants’ perspectives, striving to present an authentic and comprehensive understanding of their experiences [19]. This method included steps, including familiarization, identifying significant statements, formulating meanings, clustering themes, developing an exhaustive description, producing the fundamental structure, and seeking verification of the fundamental structure [19]. Each team member read a subset of interviews, formulating the themes after summarizing and extracting the meaningful contents, bracketing the presumptions of the researchers. This process resulted in five sub-themes, which were further categorized into two broad themes as summarized in Table 1.

**Table 1.** Emergent thematic levels.

| Themes | Sub-themes | Codes |
| --- | --- | --- |
| Culture of dating | Reasons for dating | Seeking companionship; Sharing ideas; Good company; Peer pressure; Curiosity; Need satisfaction; Pleasure |
|  | Process of partner's choice | Emotional attraction; Family-oriented goals; Physical attraction; Financial rewards; Non-financial rewards |
| Awareness and risk-taking behaviors | HIV transmission | Mother-to-Child Transmission; Unprotected sex; Soiled objects; Blood transmission; Blood pacts |
|  |  | Weight loss; itching; and diarrhea |
|  | HIV risk-taking behaviors and prevention methods | Abstinence; Sexual vagrancy; Safe sex; PrEP; Condoms; Practicing secondary abstinence; Dual protection nature |
|  | Gender roles in safe sex practice decision-making | Shared vs. individual responsibility; Prevention; HIV |
|  |  | testing |

### 2.3. Trustworthiness

To ensure the integrity and consistency of the data, known as trustworthiness, the team applied Lincoln and Goba’s evaluative criteria, encompassing credibility, transferability, dependability, and confirmability [20]. The researchers extracted and categorized text associated with the codes, comparing similarities and differences. Given that the interviewers were locals with an understanding of the regional culture, they actively participated in this analytical process. The coding tree’s dependability was affirmed through a consensus among team members on the definitions and criteria for code inclusion or exclusion. Confirmability was attained by meticulously documenting and archiving all stages of the research process, including multiple revisions and discussions of the primary and secondary categories identified by the researchers.

### 2.4. Reflexivity

In terms of reflexivity, the diverse and multinational qualitative research team, proficient in multiple languages and disciplines, worked to minimize potential biases in the analysis. The inclusion of locally trained interviewers aided in capturing cultural nuances and facilitated acceptance within the community. This aspect of the research was reflexive and collaborative, engaging continuously with the NGO partner, school and government officials, and development organizations. Despite the team’s diverse composition, potential biases in the conclusions could not be entirely ruled out. To address this, triangulation was applied, involving sharing findings with community members not interviewed and experts on the subject to identify possible inaccuracies or biases overlooked by the research team.

### 2.5. Ethical considerations

Regarding ethical considerations, approval was obtained from both the Harvard T.H. Chan School of Public Health’s Institutional Review Board (protocol #IRB21-1641) and the National Committee on Bioethics of Chad (#036CMT/PC/PMT/MESRI/SG/CNBT/2022). Informed consent forms were read aloud by the interviewers before the FGDs began. Participants had the opportunity to ask questions during the consent process and gave verbal consent. The research did not involve participants in its design, conduct, reporting, or dissemination due to practical and ethical limitations.

### 2.6. Limitation

This study included some limitations. Firstly, the study utilized descriptive phenomenology, a method that can potentially lead to researcher bias influencing everything from the research question’s development to data interpretation. To mitigate this, efforts were made to reduce the impact of the researchers’ personal beliefs, values, and experiences. This was achieved by thoroughly reviewing and discussing focus group discussion (FGD) transcripts before conducting interviews, thereby ensuring that any assumptions made by the interviewer did not overshadow the participants’ actual experiences. Secondly, our study focused solely on high school students, excluding those not in formal education, who comprise 70 to 82% of the Chadian youth population. Finally, the study did not specifically address populations most at risk for HIV, such as youth involved in sex work, MSM, transgender individuals, and injection drug users. This omission is significant considering the pivotal role these groups play in HIV transmission, where young people are not only overrepresented but also tend to have higher infection rates within these groups.

## 3. Results

Data collection included forty-eight in-classroom high school students divided into four FGDs. Each group included an equal number (n=12) of males and females. Of the 48 participants, 17 and 18 students were respectively enrolled in 10^th^ and 11^th^ grades representing the largest group (73%) of the sample (Table 2). The mean age of the participants was approximately 17 years. The IDs used in this study were not known to anyone outside of the research group.

**Table 2.** Sociodemographic characteristics of participants.

| Participant characteristics | N (%) or mean (SD) |
| --- | --- |
| **High school student sex** |  |
| Male | 24 (50%) |
| Female | 24 (50%) |
| **High school grade** |  |
| 9<sup>th</sup> grade | 6 (12.5%) |
| 10<sup>th</sup> grade | 17 (35.5%) |
| 11<sup>th</sup> grade | 18 (37.5%) |
| 12<sup>th</sup> grade | 7 (14.5%) |

By employing descriptive phenomenology, the analysis of the youth’s life experiences of supportive norms revealed two main themes: the culture of dating and risk-taking behaviors. Each theme includes sub-themes, as summarized in Table 1. Select respondent quotations, further illuminating the results shared below, can be found in Tables 3 and 4 for the dating culture and risk-taking behaviors about HIV, respectively.

**Table 3.** Selected quotes for the dating culture among the youth in Chad.

| Sub-themes | Codes | Selected quotes |
| --- | --- | --- |
| Reasons for dating | Seeking companionship | “A friend to share ideas with” (BM, #24, age range 16-20) |
|  | Felt peer-pressured | “I believe that what motivates young people like me to practice sex is peer pressure and desire” (BF, #20, age range 11-15) |
|  | Motivations driven by sexual behavior | “There were differences in the needs of young women and men” (BM, #5, age range 11-15) |
|  |  | “Need satisfaction” (NBM, #25, age range 16-20)<br>“Men of our age have sex just to satisfy themselves and for pleasure” (NBM, #28, age range 16-20) |
| Process of partners’ choice | Intrinsic motivation | “The culture around dating is focused on emotional attraction and sharing ideas” (BF, #21, age range 16-20)<br>“What is involved in dating is to...end up with a marriage around the family and having common projects” (BM, #5, age range 11-15) |
|  | Extrinsic motivation | “Our choice of partners is based on physical appearance” (BM, #3, age range 16-20)<br>“We date men because of money” (NBF, #39, age range 16-20)<br>“for me, I choose my partner based on the fact that she is helpful, and she offers me gifts” (NBM, #26, age range 16-20) |

**Table 4.**
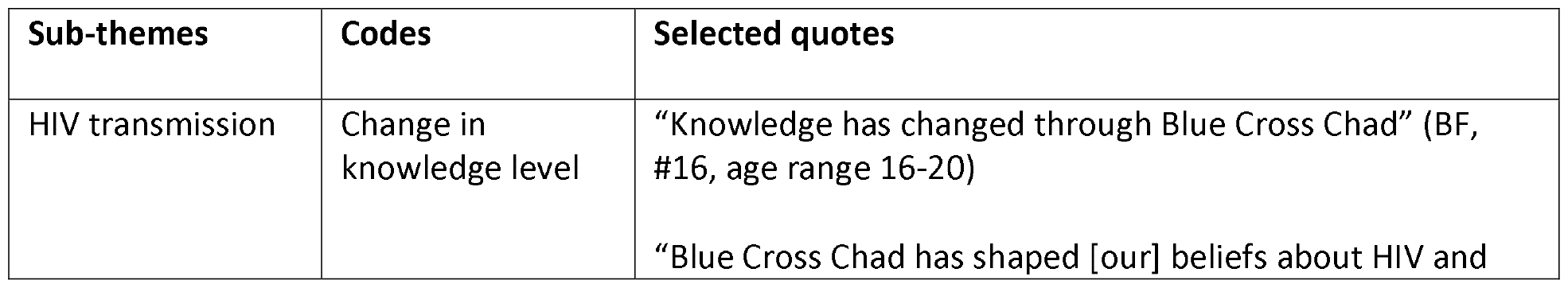
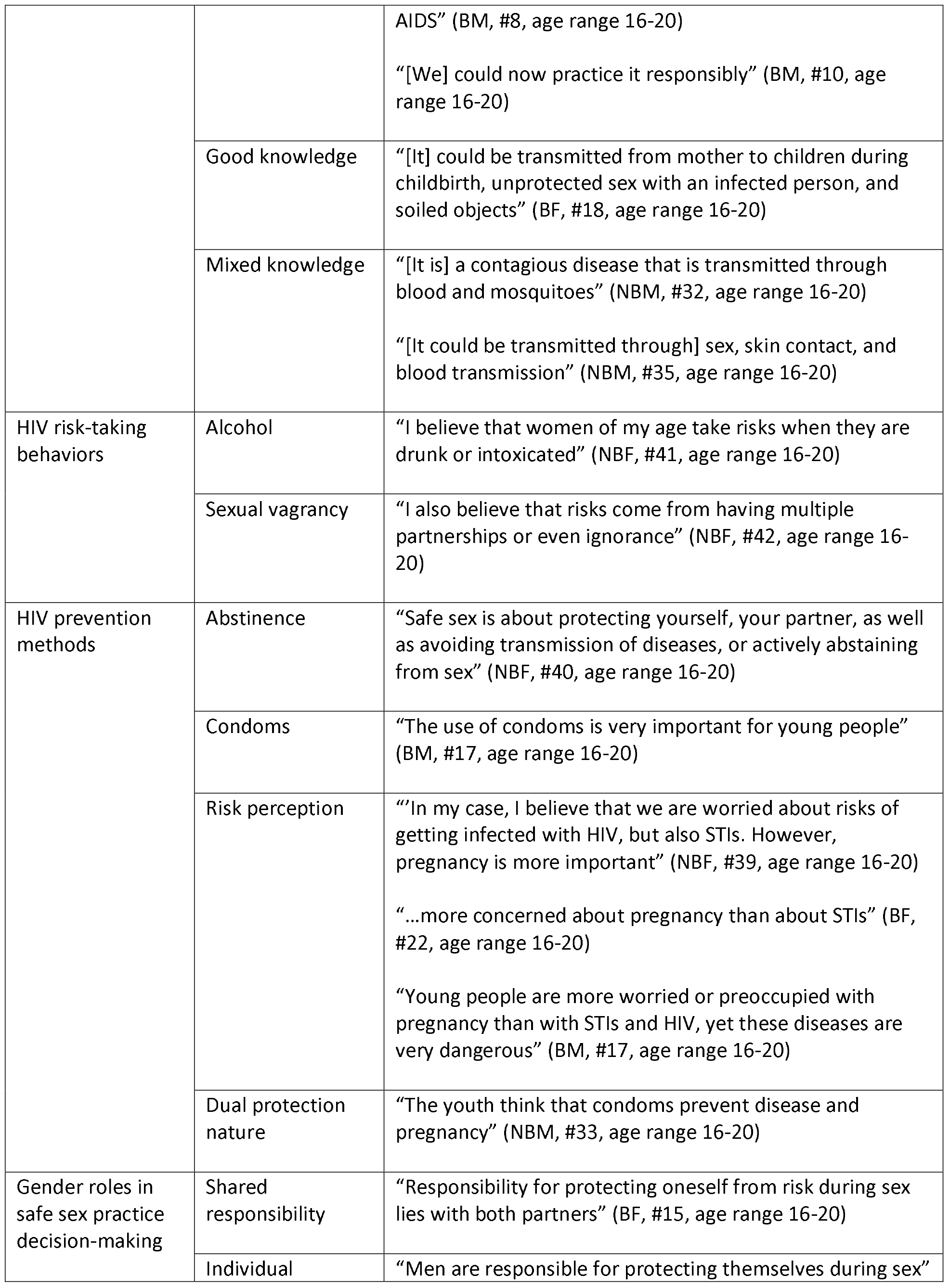
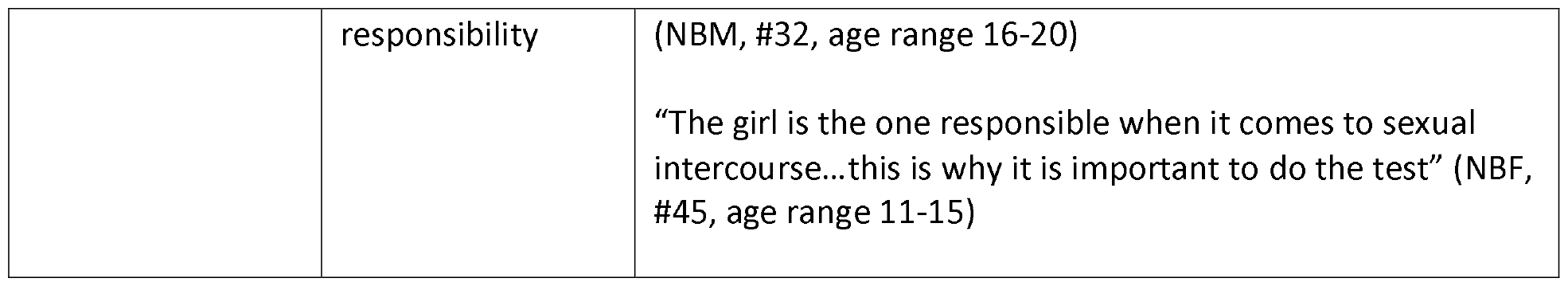
Selected quotes for risk-taking behaviors about HIV.

| Sub-themes | Codes | Selected quotes |
| --- | --- | --- |
| HIV transmission | Change in knowledge level | “Knowledge has changed through Blue Cross Chad” (BF, #16, age range 16-20)<br>“Blue Cross Chad has shaped [our] beliefs about HIV and |
|  |  | AIDS” (BM, #8, age range 16-20)<br>“[We] could now practice it responsibly” (BM, #10, age range 16-20) |
|  | Good knowledge | “[It] could be transmitted from mother to children during childbirth, unprotected sex with an infected person, and soiled objects” (BF, #18, age range 16-20) |
|  | Mixed knowledge | “[It is] a contagious disease that is transmitted through blood and mosquitoes” (NBM, #32, age range 16-20)<br>“[It could be transmitted through] sex, skin contact, and blood transmission” (NBM, #35, age range 16-20) |
| HIV risk-taking behaviors | Alcohol | “I believe that women of my age take risks when they are drunk or intoxicated” (NBF, #41, age range 16-20) |
|  | Sexual vagrancy | “I also believe that risks come from having multiple partnerships or even ignorance” (NBF, #42, age range 16-20) |
| HIV prevention methods | Abstinence | “Safe sex is about protecting yourself, your partner, as well as avoiding transmission of diseases, or actively abstaining from sex” (NBF, #40, age range 16-20) |
|  | Condoms | “The use of condoms is very important for young people” (BM, #17, age range 16-20) |
|  | Risk perception | “In my case, I believe that we are worried about risks of getting infected with HIV, but also STIs. However, pregnancy is more important” (NBF, #39, age range 16-20)<br>“...more concerned about pregnancy than about STIs” (BF, #22, age range 16-20)<br>“Young people are more worried or preoccupied with pregnancy than with STIs and HIV, yet these diseases are very dangerous” (BM, #17, age range 16-20) |
|  | Dual protection nature | “The youth think that condoms prevent disease and pregnancy” (NBM, #33, age range 16-20) |
| Gender roles in safe sex practice decision-making | Shared responsibility | “Responsibility for protecting oneself from risk during sex lies with both partners” (BF, #15, age range 16-20) |
|  | Individual | “Men are responsible for protecting themselves during sex” |
|  | responsibility | (NBM, #32, age range 16-20)<br>“The girl is the one responsible when it comes to sexual intercourse...this is why it is important to do the test” (NBF, #45, age range 11-15) |

### 3.1. Taking a step back: What is the culture of dating among the youth in Chad?

#### 3.1.1. Reasons for dating

Students expressed various reasons for dating; however, two main categories emerged, including those who were in a relationship because they were seeking companionship and others whose motivations were driven by sexual behavior. Among females, the search for companionship was shared as the primary reason for dating. Although their initial reason for dating was due to wanting good company, some female beneficiaries also indicated they felt pressured by their partners to engage in sex during their relationship. On the other hand, the interest in dating was primarily driven by sex for male beneficiaries. Males recognized that their interests in dating were different from their female counterparts. They mentioned that it was due to their “*need satisfaction*” (NBM, #25, age range 16-20) and further elaborated by stating that *“this means that for us, sex is a sign of maturity, or it means that we want to test our masculinity*.*”* (BM, #11, age range 16-20)

#### 3.1.2. The process of partners’ choice

In interviews, there were multiple reasons discussed for choosing a specific partner. Partners’ choices were classified into two main categories: intrinsic and extrinsic motivations, as summarized in Table 3. In this study, emotional attraction remained one of the reasons for choosing a partner for both females and males. Additionally, male beneficiaries stated that their partner choice depended on their motivation to form a family.

As mentioned above, extrinsic motivations were also identified. First, physical attraction was mentioned as a motivation for males and females, whether it was between and within groups. In addition, there were other extrinsic motivations, more specifically being motivated by financial rewards, such as money and other financial reasons.

### 3.2. Awareness and risk-taking behaviors about HIV

The previous section dissected the nuances of dating culture among the youth in Chad, analyzing their motivations, behaviors, and the social contexts of their romantic relationships. The following section presents a detailed examination of the youth’s understanding and management of HIV risks, elucidating the critical connection between their dating lives and vulnerability to HIV.

#### 3.2.1. HIV transmission

Overall, the beneficiary groups highlighted a change in their level of knowledge post-peer education program in their respective high schools. They further stated that this latter has left them feeling more prepared when having sex. Moreover, males and females in the beneficiary group, as well as females in the non-beneficiary group, were able to correctly explain what the disease entailed, its transmission routes, and associated symptoms. They frequently mentioned perinatal (mother-to-child), or unprotected sex with an HIV infected person as HIV transmission modes. However, males in the non-beneficiary group showed some mixed knowledge.

#### 3.2.2. HIV risk-taking behaviors

When it comes to risky sexual behaviors and HIV, all groups identified several factors that may place individuals at greater risk. Alcohol was indicated as a potential factor. Some also identified multiple concurrent partnerships.

#### 3.2.3. HIV prevention methods

Both female beneficiaries and non-beneficiaries mentioned that abstinence was a popular prevention method. On the other hand, male beneficiaries highlighted that abstaining is *“abstaining is hardly encouraged among them”* (BM, #8, age range 16-20) since most of them are sexually active at a young age.

Condoms were identified as an important prevention method among the youth. Even though the use of condoms was commonly cited to practice safe sex, it was primarily seen as pregnancy prevention and not to reduce the risk of HIV or other STIs in both female beneficiaries and non-beneficiaries. On the other hand, this was only cited in beneficiary males who recognized a higher risk perception for pregnancy than for STIs among Chadian youth. For this reason, condoms were attractive to them because of their dual protection nature.

#### 3.2.4. Gender roles in safe sex practice decision-making

In the beneficiary group, both males and females believed that they had a shared responsibility when it came to practicing safe sex. This shared responsibility was not identified in the non-beneficiary group and differed across gender. In the non-beneficiary male group, it was noted that the responsibility of men lay in only wearing a condom, whereas women were responsible for getting tested for HIV.

## 4. Discussion

We conducted a qualitative study to delve into the perceptions and experiences of Chadian youth regarding HIV supportive norms, comparing students who received peer HIV education with those who did not. First, our findings revealed distinct motivations for dating among Chadian youth: females primarily sought companionship, while males were driven by sexual desires, often leading to pressure for sex in relationships. Partner choice hinged on emotional and physical attraction, with extrinsic motivations like financial benefits also playing a role. Post-education, both genders showed improved HIV knowledge, identifying transmission routes and prevention methods. However, misconceptions persisted, especially among non-beneficiary males. Risky behaviors like alcohol use and multiple partnerships were recognized, and while condom use is common, it was seen more for pregnancy prevention than for HIV/STI protection. Finally, gender differences emerged in perceptions of safe sex practices, with beneficiary groups viewing it as a shared responsibility, unlike non-beneficiaries.

Our findings revealed distinct reasons for dating among Chadian youth, especially males and females. The search for companionship identified among female beneficiaries and sexual satisfaction in males were also observed in a study (2007) conducted among inner-city black adolescents in the United States [21]. According to Andrinopoulos et al. (2007), young women often look for romantic connections, whereas young men pursue both sexual and romantic relationships [21]. These findings were in line with Buss’ sexual strategies theory, which suggests distinct approaches to sexual relationships by men and women [22]. Additionally, the concept of sexual vagrancy, described by men as a way to prove their masculinity, was a common social phenomenon among young men. This notion was supported by Andrinopoulos et al. (2007), who noted that having multiple partners was seen as a means for young men to elevate their social standing among peers [21].

Our study identified two primary categories in partner selection criteria: intrinsic and extrinsic motivations. Intrinsic motivation, stemming from internal characteristics such as personality, values, and interests, was evident in the preference for emotional connection and a family-centric partnership [23]. This reflected a deep, value-driven approach to relationships, highlighting the importance of personal and emotional factors in choosing a partner. On the other hand, extrinsic motivation is influenced by external factors like physical attractiveness and financial status. The focus on these superficial aspects suggests that physical and material attributes played a significant role in partner selection. This dichotomy in motivations enhanced our understanding of partner selection, especially among Chadian youth, providing a comprehensive perspective on the interplay between deep-seated values and external allurements in choosing a partner. Further research in various African contexts supported these findings. A study in Ghana by Ampofo (1997) among young women showed that love, affection, and the desire for children were crucial in partner selection, aligning with the intrinsic factors we identified [24]. However, financial capability was also a significant consideration, resonating with our observations of extrinsic motivations [24]. This economic aspect of partner selection was noted in Meekers and Calvès’ study (1992) in Cameroon, particularly among women in low-income sectors who viewed financial support as a means of survival or accessing capital [25]. Similarly, Nicole De Wet’s research in South Africa (2019) found that young women, especially those aged 20-24 and students, were more likely to choose older partners for financial benefits [26]. This underscored the complex interplay of emotional, personal, and economic factors in partner selection across different African cultures.

A concerning revelation from our study is the gap in HIV awareness, particularly among non-beneficiary males. This group displayed misconceptions about HIV transmission, underscoring the need for targeted educational interventions. However, increased knowledge did not necessarily translate into safer behaviors, as seen in persistent risky practices like intoxication and unprotected sex. This echoed Swahn et al.’s findings (2021) on the overlooked roles of alcohol consumption and gender-based violence in HIV risk [27]. Furthermore, our findings highlighted that young women often feel pressured into sexual activities, which were in line with Plichta et al.’s observations (1991) about the lack of control women have over sexual encounters and condom use [28]. This latter was also confirmed by Duby et al. (2021), especially mentioning the power inequities in condomless sex for South African adolescent girls and young women [29]. The gendered attitudes towards sexual health responsibility, shaped by societal norms, call for gender-sexual health education. Studies like Wathuta (2016) emphasize the need for accurate gender dynamics representation in HIV responses [30]. Moreover, Ruppe et al. (2021) highlighted the role of women’s healthcare providers in HIV prevention resonating with our findings in Chad [31].

The absence of awareness about Pre-Exposure Prophylaxis (PrEP) in Chad is a significant gap in HIV prevention. As PrEP is not readily available in Chad, advocacy efforts are crucial for its accessibility. Despite this, there’s a pressing need to start raising awareness about PrEP’s effectiveness in HIV prevention. Current prevention strategies primarily focus on abstinence and condom use, driven more by pregnancy concerns than HIV or STIs, especially among females. This reflects the societal prioritization of certain risks over others. However, the prevalent ‘abstinence’ culture contrasts sharply with the reality of adolescent premarital sex, often leading to stigma rather than open discussion [32, 33]. Additionally, the use of abstinence as a strategy for preventing HIV has been found to be ineffective [32, 33]. Addressing this mismatch requires a shift in societal norms. Engaging adolescents, families, educators, and community leaders is crucial in this endeavor. Peer health education approaches have shown promise in shifting attitudes toward sexual and reproductive health, but their design needs careful consideration to address these complex issues effectively.

## 5. Conclusions

The manuscript provided a comprehensive look at the complex interplay between adolescent dating behaviors and HIV perceptions among Chadian youth, highlighting the cultural and social influences on youth attitudes towards dating and HIV. Significant gender-based differences in dating motivations and partner selection criteria, as well as disparity in HIV knowledge and prevention awareness among adolescents were uncovered. This study underscores the need for targeted, gender-sensitive educational interventions. Notably, there was an absence of awareness about PrEP in Chad, signaling a gap in HIV prevention strategies, which are currently more focused on abstinence and condom use, driven largely by concerns over pregnancy rather than HIV or STIs. Future research should focus on addressing the realities of adolescent sexual behavior, emphasizing the role of community stakeholders in fostering effective and comprehensive sexual health education and prevention strategies. This approach will enrich the literature with dynamic insights and practical implications for HIV prevention among Chadian youth.

## Data Availability

The availability of the full data set is not publicly available in a repository due to ethical restrictions. Ethical approval for the study was received from both the Harvard T.H. Chan School of Public Health’s Institutional Review Board (protocol #IRB21-1641) and the National Committee on Bioethics of Chad (#036CMT/PC/PMT/MESRI/SG/CNBT/2022). When applying for ethical approval we did not specify that the data would be made publicly available in a repository. As part of the written and verbal consent we assured participants that all data would be confidential and access to the recordings would be restricted to the research team. We did specify that “some of their words” may be used in reporting the findings of the study (which we have done within the manuscript as non-identifiable quotes), however to make all raw data publicly available will be a serious breach to the rights of ethical of participants given did not consent to this.

## Author Contributions

**Conceptualization:** Esias Bedingar, Christopher Sudfeld.

**Methodology:** Esias Bedingar, Ngarossorang Bedingar, Christopher Sudfled.

**Investigation:** Esias Bedingar, Ngarossorang Bedingar.

**Data curation:** Esias Bedingar, Ngarossorong Bedingar.

**Formal analysis:** Esias Bedingar.

**Funding acquisition:** Esias Bedingar.

**Project administration:** Esias Bedingar, Ngarossorang Bedingar.

**Supervision:** Esias Bedingar.

**Visualization:** Esias Bedingar.

**Writing – original draft:** Esias Bedingar.

**Writing – review & editing:** Esias Bedingar, Djimet Seli, Christopher Sudfeld.

## Acknowledgements

We are grateful for the collaboration and support from Blue Cross Chad from which this research originated. We thank the participants for their time and willingness to share their experiences with us.

## Supporting information

S1 Appendix: Semi-structured Focus Group Discussion Topic Guide

S2 Appendix: COREQ Checklist

